# Planning for the aftershocks: a model of post-acute care needs for hospitalized COVID-19 patients

**DOI:** 10.1101/2020.06.12.20129551

**Authors:** Matthew Maloney, Ryan Morley, Robert Checketts, Peter Weir, Darryl Barfuss, Hannah R. Meredith, Michelle G. Hofmann, Matthew H. Samore, Lindsay T. Keegan

## Abstract

Since its emergence in late 2019, COVID-19 has caused significant global morbidity and mortality, overwhelming health systems. Considerable attention has been paid to the burden COVID-19 has put on acute care hospitals, with numerous models projecting hospitalizations and ICU needs for the duration of the pandemic. However, less attention has been paid to where these patients may go if they require additional care following hospital discharge. As COVID-19 patients recover from severe infections, many of them require additional care. Yet with post-acute care facilities averaging 85% capacity prior to the pandemic and the significant potential for outbreaks, consideration of the downstream effects of the surge of hospitalized COVID-19 patients is critical. Here, we present a method for projecting COVID-19 post-acute care needs. Our model is designed to take the output from any of the numerous epidemiological models (hospital discharges) and estimate the flow of patients to post-acute care services, thus providing a similar surge planning model for post-acute care services. Using data from the University of Utah Hospital, we find that for those who require specialized post-acute care, the majority require either home health care or skilled nursing facilities. Likewise, we find the expected peak in post-acute care occurs about two weeks after the expected peak for acute care hospitalizations, a result of the duration of hospitalization. This short delay between acute care and post-acute care surges highlights the importance of considering the organization necessary to accommodate the influx of recovering COVID-19 patients and protect non-COVID-19 patients prior to the peak in acute care hospitalizations. We developed this model to guide policymakers in addressing the “aftershocks” of discharged patients requiring further supportive care; while we only show the outcomes for discharges based on preliminary data from the University of Utah Hospital, we suggest alternative uses for our model including adapting it to explore potential alternative strategies for addressing the surge in acute care facilities during future pandemic waves.

**Author Summary:** COVID-19 has caused significant morbidity and mortality globally, putting considerable strain on healthcare systems as a result of high rates of hospitalization and critical care needs among COVID-19 patients. To address this immediate need, a number of decision support tools have been developed to project hospitalization, intensive care unit (ICU) hospitalizations, and ventilator needs for the COVID-19 pandemic. As COVID-19 patients are discharged from acute care hospitals, many of them will require significant additional post-acute care. However, with post-acute care facilities at high capacity prior to the influx of COVID-19 patients and with significant outbreak potential in long-term care facilities, there is high potential for shortages of post-acute care services. Here, we present a model of COVID-19 post-acute care needs that is analogous to most epidemiological models of COVID-19 hospitalization and ICU care needs. We develop our model on University of Utah Hospital data and demonstrate its utility and its flexibility to be used in other contexts. Our model aims to guide public health policymaking in addressing the “aftershocks” of discharged patients requiring further care, to prevent potential healthcare shortages.

## Introduction

Since its emergence in late 2019, the virus responsible for coronavirus disease 2019 (COVID-19) has spread rapidly, causing significant global morbidity and mortality [1,2]. While COVID-19 has considerable individual health impacts, significant attention has been paid to the strain put on health systems resulting from high rates of hospitalization, critical care, and ventilation among COVID-19 infections [3,4]. Consequently, a large focus of decision support tools, including epidemiological models, has been to project the impact of COVID-19 on acute care hospitalizations and ICU admissions under different non-pharmaceutical interventions to help guide public health action.

Epidemiological models have played a signficant role in shaping the public health response globally. A wide variety of methods have been deployed to project the course of the COVID-19 outbreak, including agent-based models, population-level models with and without age structure, and curve fitting approaches [5–9]. Despite the variation in methodology, they each estimate the same health outcomes: infections, hospitalizations, ICU hospitalizations, ventilators needed, and deaths. These models played a critical role in helping policy makers design and implement effective interventions to avoid the universal projection that without interventions, the surge of COVID-19 patients at the peak of the outbreak would overwhelm healthcare facilities. While these health outcomes provide necessary projections to address the surge of patients expected during local epidemic peaks, none of these approaches consider what could be described as the “aftershocks” of the surge. That is, a secondary surge due to patients who were discharged from an acute care facility, but still require continued support.

As patients recover from severe or critical COVID-19 infections, many will require rehabilitative, supportive, or palliative care services. Depending on the intensity of care the patient requires, treatment might include a stay in a skilled nursing facility or care at home, including hospice care. Additionally, post-acute care services can act as a pressure release valve for hospitals reaching saturation [10]. Grabowski et al. [10] highlights some of the challenges for post-acute care planning. As evidenced by COVID-19 outbreaks in nursing facilities across the globe [11], skilled nursing facilities are ill-equipped to manage the infection prevention and control measures essential to containing the spread of COVID-19. Skilled nursing facilities are, on average, at 85% capacity [12], and no single facility is likely prepared to handle a surge of new patients. Any given hospital discharges patients to multiple facilities. Since these recovering COVID-19 patients may still be infectious, they further increase the already high risk for outbreak at these facilities and, subsequently, the need for even more patients requiring acute hospital care. The staggering number of excess deaths attributed to COVID-19 [13,14] originating from nursing facilities is a testament to the critical need for surge planning modeling that integrates acute hospital and post-acute care.

We consider the downstream effects on post-acute care services of COVID-19 patients who previously required hopitalization, ICU care, and/or mechanical ventilation. In this paper, we present a method for projecting post-acute care capacity needs that takes in a time series of discharge estimates from any of the numerous epidemiological models of COVID-19 and extends the results from these models to estimate the flow of patients to three post-acute care services (and direct-to-home), thus providing the same type of surge planning model for post-acute care services that the multitude of high-profile epidemiological models provide for acute care planning.

## Methods

This model is designed to extend projections from existing epidemiological models. It builds off of the standard model outputs (time series of hospital and ICU discharges) and projects the post-acute care needs of discharged patients. Post-acute care services include: 1) direct-to-home (none), 2) home health care (hh), 3) skilled nursing facility (snf), and 4) hospice (hos). This model is available in the Github repository “mattrmaloney/covid-post-acute-care.” [15]

### Model of discharge allocation

To project patient movement to each of the four post-acute care services, we use the following statistical model, which uses a multinomial distribution to estimate patient flow. We estimate the discharge location for *n*_*j*_ patients discharged from ward *j*, where *j* is either patients in the hospital for COVID-19 (all wards) or patients in the ICU:

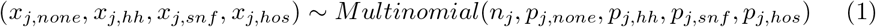

where *p*_*j,k*_ and *x*_*j,k*_ denotes the probability and number, respectively, of patients discharged from ward *j* to post-acute outcome *k* (where *k* ∈ *{*none, *hh, snf, hosp*} for our model). Thus, we have that ∑_*k*_ *x*_*j,k*_ = *n*_*j*_. In addition, the set of probability parameters must sum to one, so for our model *p*_*j,none*_ + *p*_*j,hh*_ + *p*_*j,snf*_ + *p*_*j,hos*_ = 1.

The multinomial patient allocation function is sufficient to determine flows into post-acute care services. Formally, the model takes a time series of hospital discharges from ward type *j*, i.e.

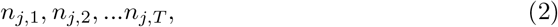

where *T* denotes the total length of the time series. At each time step, a draw from the multinomial distribution is used to allocate discharge counts across post-acute care services. The result is a matrix with dimensions *T* × 4, where each column corresponds to the number of new patients that flow to each post-acute care service at each point in time.

To further operationalize this model, we also estimate the number of patients in each of these services at any time to provide guidance on when existing services may reach or exceed capacity. To determine patient counts in each service, we require baseline estimates of the number of patients in each service and the length of stay for patients in each service. In this paper, we assume the initial patient census is zero, however this assumption can be easily changed. In the next section, we detail how length of stay estimates were determined. To calculate the census over time for each service, we use the length-of-stay estimates to create discharge series for each service. The number of discharges each day is equal to the inflow from *l*_*k*_ days ago, where *l*_*k*_ is the length-of-stay assumption for service *k*. The cumulative sum of the inpatient flows minus the cumulative sum of discharges gives the census at each point in time.

## Model inputs and parameters

### Hospital and ICU discharges

Since our model does not attempt to model the broader COVID-19 outbreak, it requires a time series of hospital and ICU discharges. In this paper, we use model outputs from the Johns Hopkins University Infectious Disease Dynamics (JHU IDD) model, which combines an SEIR model with a statistical model to estimate daily hospitalizations, ICU hospitalizations, and ventilated patients [8]. As a result of the considerable uncertainty surrounding estimates of the proportion of all infections that are hospitalized, the JHU IDD model assumes the hospitalization rate is 10 times the infection fatality rate (IFR) and, due to uncertainty in estimates of the IFR, they explore 3 IFRs: 0.25%, 0.5%, and 1%. Here, we explore 0.5% IFR and the corresponding 5% hospitalization rate.

We demonstrate the utility of our model on two example runs of the COVID-19 outbreak in Utah using the JHU IDD model, simulating the outbreak with *R*_*t*_ = 1.2 for “current Utah”, based off of the actual transmission rate in Utah on May 2,2020, calculated based off of the positive test rates [16]; and *R*_*t*_ = 1.6 followed by pulsed lockdowns with *R*_*t*_ = 0.55 for “pulsed social distancing”, calculated based of Maryland’s pre-lockdown reproductive number and the estimate of the lockdown in the UK [16,17]; for both we assume a 0.5% IFR. We run the model from January 1, 2020 to December 31, 2020 (Figure 1).

**Fig 1.**
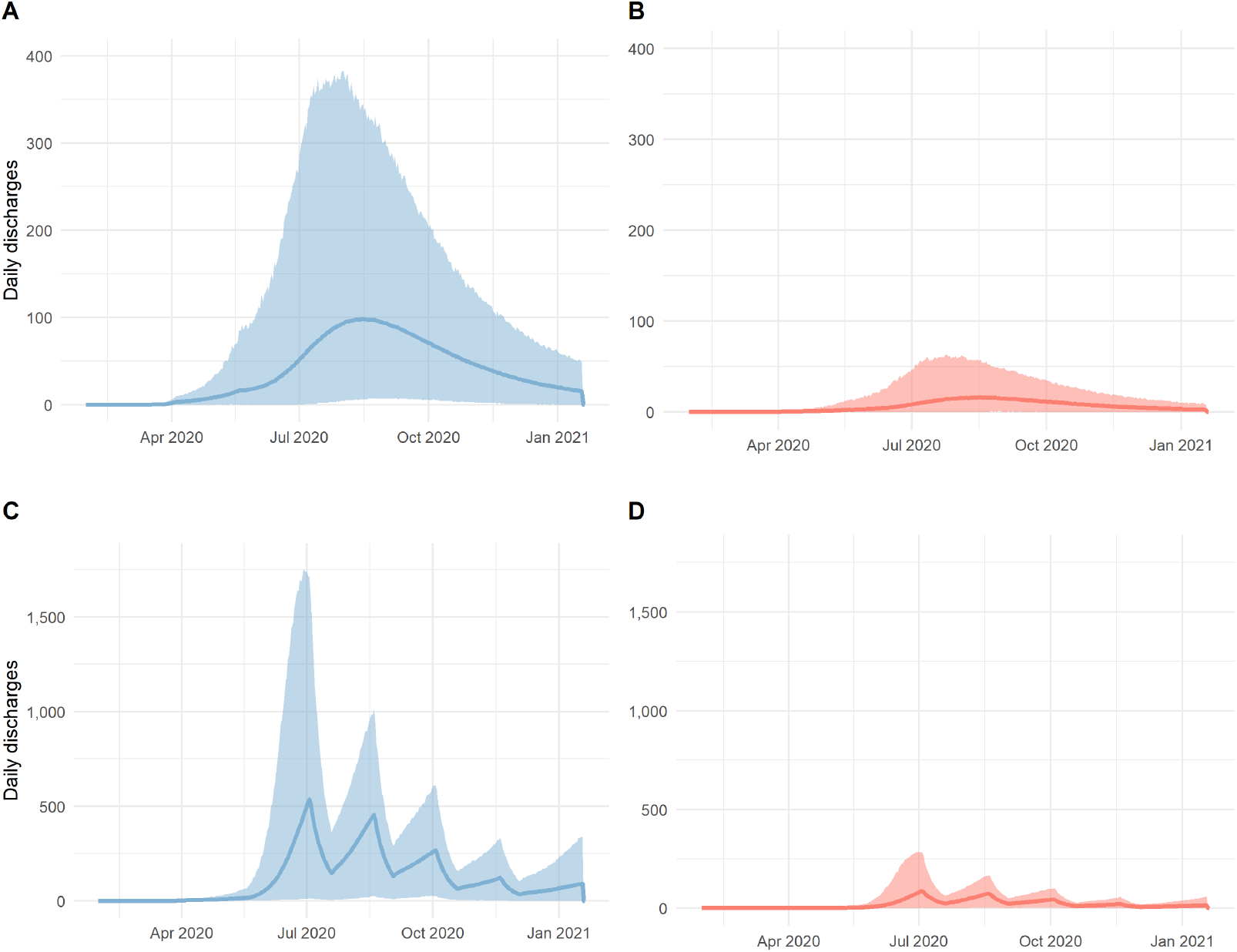
Timeseries of hospital and ICU A) discharges from hospital across the state of Utah based on an epidemiological model of the current reproductive number in Utah (current Utah); B) discharges from ICUs across the state of Utah based on an epidemiological model of (current Utah); C) discharges from the hospital across the state of Utah based on an epidemiological model of pulsed lockdowns (opening based on ICU capacity); and D) discharges from ICUs across the state of Utah based on an epidemiological model of opening based on ICU capacity

### Length of stay estimates

A critical component to determining when current capacity will be exceeded by COVID-19 patient needs is estimating the duration of stay for patients who are discharged to each of the different service types (Table 1). We do not calculate a length of stay for patients discharged directly to home since that will not influence facility capacity. The estimate for the average length of stay (ALOS) for COVID-19 patients in skilled nursing facilities is based on the University of Utah Health’s ALOS from reporting affiliate skilled nursing facilities, and further validated by the actual experience in a COVID-19 dedicated skilled nursing facility in Utah after seven weeks of operations during the pandemic. The home health and hospice ALOS assumptions are based on anticipated ALOS and a small COVID-19 patient sample from the University of Utah Health’s home health and hospice agency, Community Nursing Services. The hospice ALOS also includes general inpatient hospice patients.

**Table 1:** Assumptions for length of stay and initial patient counts for post-acute care services

|  | Days |
| --- | --- |
| Direct-to-Home | 0 |
| Home Health | 15 |
| Skilled Nursing Facility | 18 |
| Hospice | 7 |

### Post-acute care probability priors

The probabilities that parameterize the multinomial distribution are very uncertain. Even as the COVID-19 pandemic has progressed with significant numbers of hospitalizations and deaths, post-acute care needs lag behind. Considering the generation time of infections coupled with the long duration of hospitalizations and ICU hospitalizations, the number of patients who have been discharged to post-acute care services remains relatively low, leading to uncertainty in parameter estimates. First, we determined preliminary estimates of the fraction of individuals who would end up in each post-acute care service type, for both non-ICU discharges and ICU discharges. Then, we constructed discrete “low,” “mean,” and “high” estimates to summarize the potential range of outcomes (Table 2).

**Table 2:** Preliminary estimated ranges for fractions of patients discharged to each post-acute care destination for non-ICU and ICU hospitalized COVID-19 patients

|  | Non-ICU discharge |  |  | ICU discharge |  |  |
| --- | --- | --- | --- | --- | --- | --- |
|  | Low | Mean | High | Low | Mean | High |
| Direct-to-Home | 0.496 | 0.748 | 0.874 | 0.152 | 0.488 | 0.744 |
| Home Health | 0.090 | 0.180 | 0.360 | 0.100 | 0.200 | 0.400 |
| SNF | 0.030 | 0.060 | 0.120 | 0.150 | 0.300 | 0.400 |
| Hospice | 0.006 | 0.012 | 0.048 | 0.006 | 0.012 | 0.024 |

The non-ICU discharge destination fractions are based on internal University of Utah Health system discharge data from the previous calendar year. The mean ICU patient discharge fractions for home health and skilled nursing facility care are based on historical estimates for those hospitalized for sepsis, which has a similar inpatient mortality rate [10]. The other ICU estimates are also based on University of Utah discharge data from the previous calendar year.

Rather than using these low, mean, and high estimates directly, we create a continuous distribution of possible multinomial parameter values using a Dirichlet distribution. Formally,

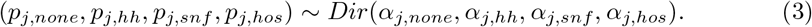

Thus, for every hospitalization discharge series, we draw a random set of probabilities/parameters for the multinomial distribution from that Dirichlet distribution, where the *α* values are parameters that will initially be proportional to the mean care service fractions shown in Table 2. Denoting the vector of these mean values for ward *j* as 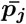, we introduce a scaling parameter, *ϕ*_*j*_, such that our initial *α* parameters are

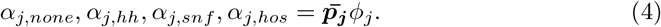

The magnitude of *ϕ*_*j*_ will determine how much weight is put on our priors versus the new, actual discharge outcomes we may observe in the future. Smaller *α* values correspond to wider, flatter distributions around each service type probability and represent less confidence in our priors about these probabilities. If we restrict *ϕ*_*j*_ to be an integer it has a nice interpretation: we are as confident in our priors as if we had observed *ϕ*_*j*_ discharges that were distributed according to 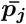.

We used the low and high estimates from Table 2 to calibrate *ϕ*_*j*_ for *j* = *icu* and *j* = *non_icu* discharges. We ran simulations to determine numerically the lowest integer values of *ϕ* at which 90% of the individual values drawn from the Dirichlet distribution were within their respective low to high ranges. We do not require that 90% of the draws of each parameter fall within their low-to-high range, only that collectively 90% of all the parameters are within their respective ranges. The former would be too restrictive. Specifically, the algorithm starts at *ϕ*_*j*_ = 1, takes 10,000 draws from the Dirichlet distribution, and then checks what percentage of the parameters fall within their low-to-high ranges. If the percentage is less than our 90% condition, we add one to *ϕ*_*j*_ and run another 10,000 simulations. We repeat the process until our condition is met. Following this procedure, we find that *ϕ*_*icu*_ = 138 and *ϕ*_*non_icu*_ = 121. The corresponding Dirichlet parameter values are shown in Table 3. The corresponding Dirichlet distributions are shown in Figure 2.

**Table 3:**
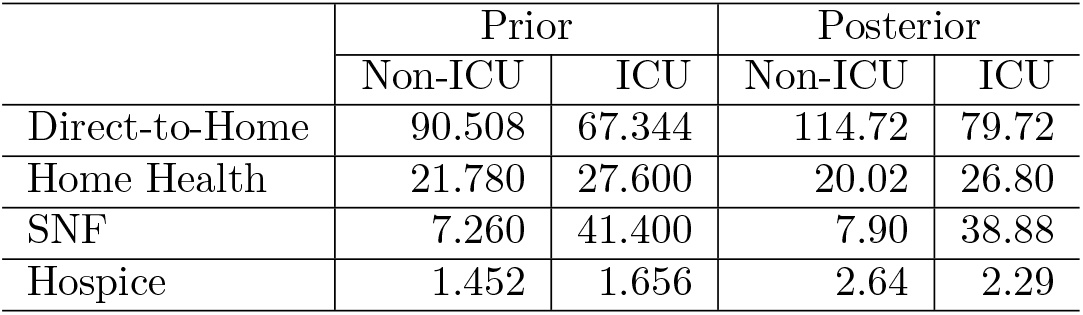
Dirichlet distribution parameters

**Fig 2.**
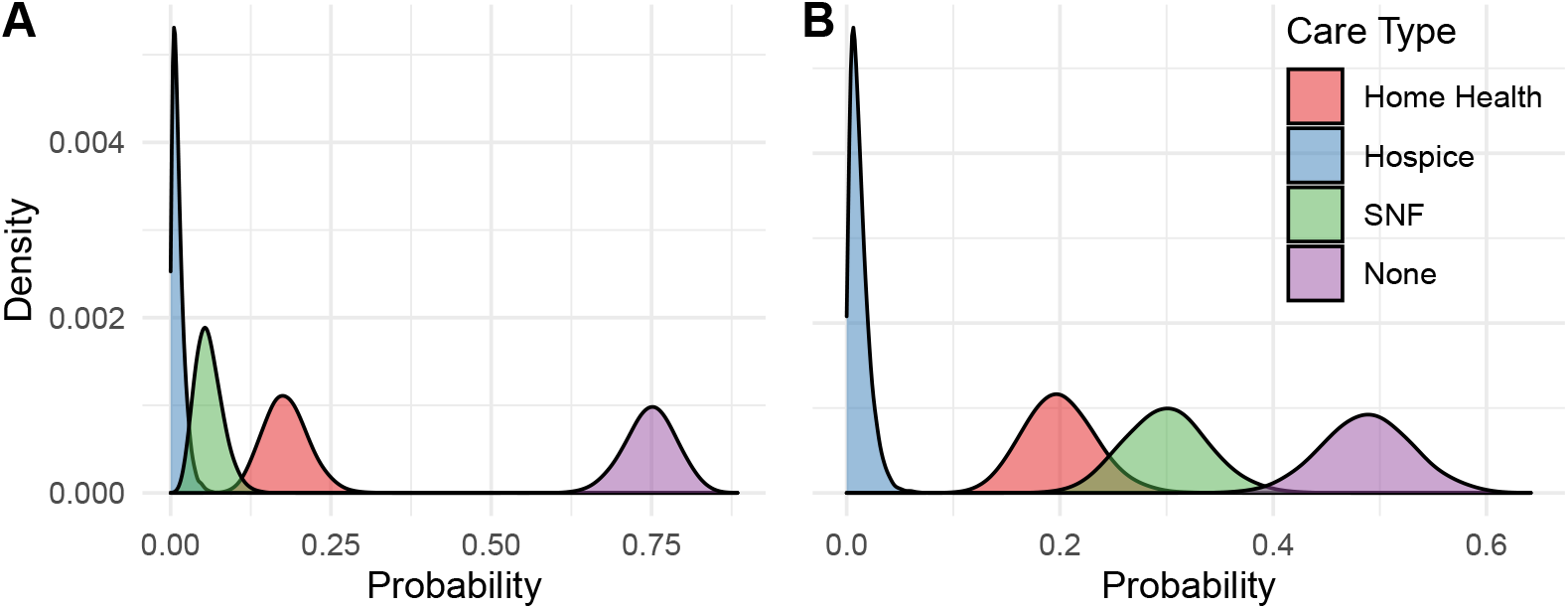
Post-acute care priors probability for each of the four post-discharge care services, (red) Home Health, (blue) Hospice, (green) Skilled Nursing Facility, (purple) None for A) non-ICU hospitalized and B) ICU hospitalized COVID-19 patients.

### Method for updating priors

As new patient counts for each care category are observed (denoted by *x*) we can incorporate this new information to update the Dirichlet distribution according to

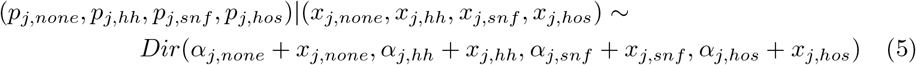

The expected probabilities in the posterior distributions are

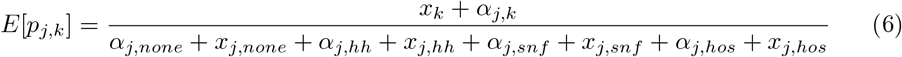

## Results

### Updating priors using University of Utah Hospital discharges

As of May 27, 2020, University of Utah Hospital has discharged a total of 78 COVID-19 patients: 24 patients who spent time in the ICU and 54 who had not. The cumulative fraction of patients who were discharged to the four post-acute care service types (as well as a dotted line indicating our initial expectations of where patients would be discharged to) are shown in Figure 3 for non-ICU patients (Figure 3A) and patients who spent time in the ICU (Figure 3B). These observations of actual patient discharges were used to update our priors. For non-ICU discharges, the observations have decreased our expectation of the percentage of patients who will require home health services and increased our expectation about the percentage of patients who will require hospice care. For ICU patients, the observations have increased our expectation of the percentage of patients who will not require any services and decreased our expectation of the percentage patients who will require a skilled nursing facility. Table 3 shows the parameters for the posterior Dirichlet distributions.

**Fig 3.**
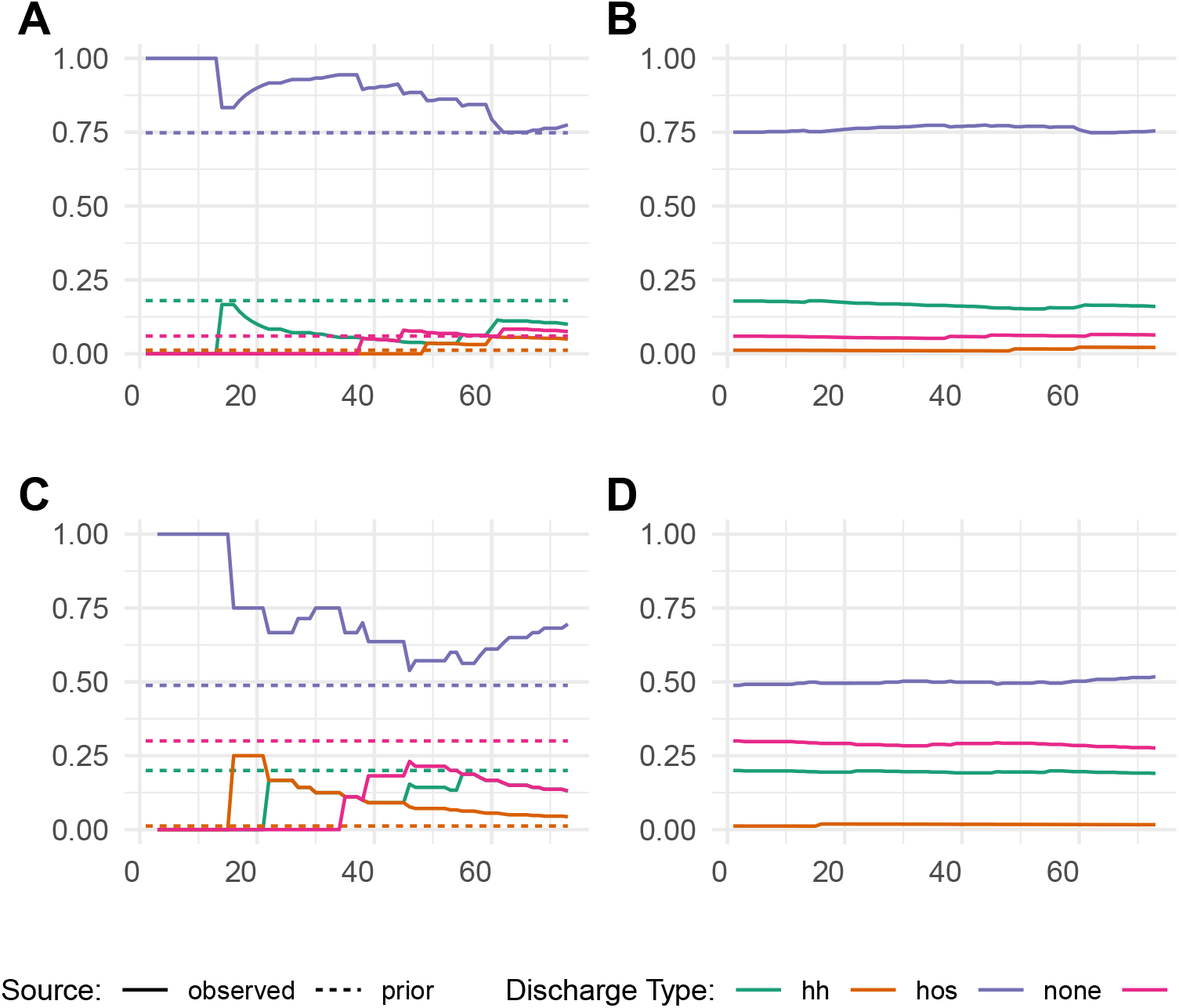
Incorporating new information over time. The x-axis shows days since we began tracking discharge data. The left-side plots compare the cumulative observed fraction of University of Utah COVID-19 patients discharged to each care service with our initial expectations for Non-ICU (A) and ICU (C) discharges. The right-side plots show how our discharge probability expectations changed over time based the observed discharges for non-ICU (B) and ICU (D).

### Post-acute care simulations

We present an example run of 1000 model simulations across the state of Utah and in Salt Lake City to demonstrate the post-acute care model output from January 31 to December 31, 2020. We ran the simulations on the hospital discharge time series shown in Figure 1.

We calculated daily estimates of both the number of patients discharged to each care type and the number of COVID-19 beds needed. Additionally, while we do not consider a baseline occupancy in this paper, one could be used to estimate the total patient census for each facility type at a given spatial scale.

We show the model ouput, a time series of the daily number of post-acute care beds needed for each of the three care types considered (home health, hospice, and skilled nursing facilities), for the State of Utah for patients discharged from all hospital wards (Figure 4) and for patients discharged from the ICU (Figure 5). Here, we find the majority of patients in Utah needing additional care are requiring home health care or skilled nursing facilities, with very few needing hospice care. Likewise, we find the expected peak in post-acute care needs occurs about two weeks after the expected peak for acute-care hospitalizations (Figure 6). This is a result of the duration of hospitalization delaying the peak of post-acute care needs. Additionally, due to the long duration of care patients receiving post-acute services require, the distribution of occupancy is asymmetric with a high volume of patients needing post-acute care services through the remainder of the year and into next year.

**Fig 4.**
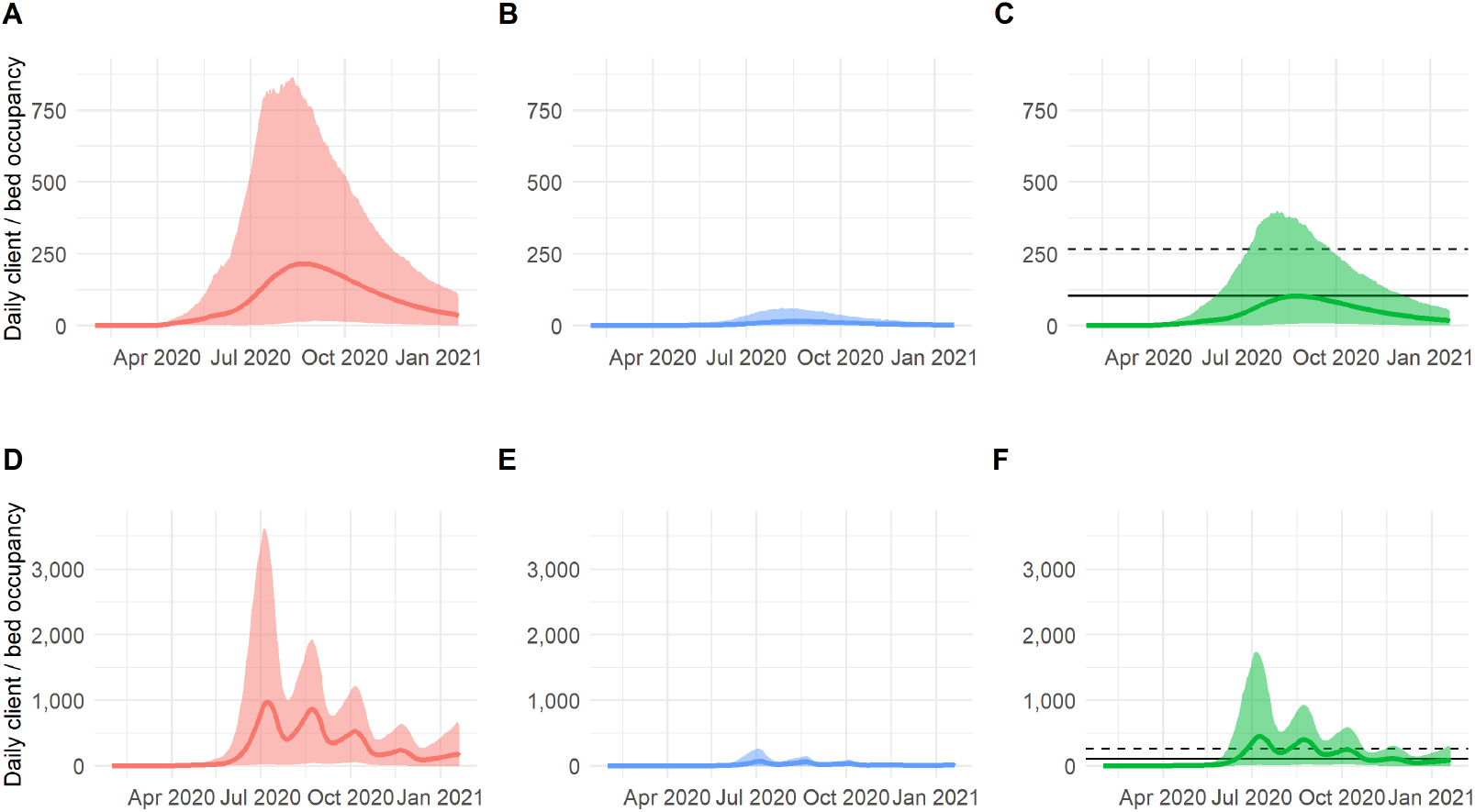
Example post-acute care daily bed occupancy needs and available skilled nursing facility beds in COVID-19 dedicated facilities (solid black line) and beds in COVID-19 dedicated wards within a larger facility (dotted black line) in the State of Utah for patients discharged from the hospital to each of three care types under two non-pharmaceutical interventions A) Home health care based on current Utah, B) Hospice care based on current Utah, and C) Skilled Nursing Facilities based on current Utah, D) Home health care based on pulsed social distancing, E) Hospice care based on pulsed social distancing, and F) Skilled Nursing Facilities based on pulsed social distancing.

**Fig 5.**
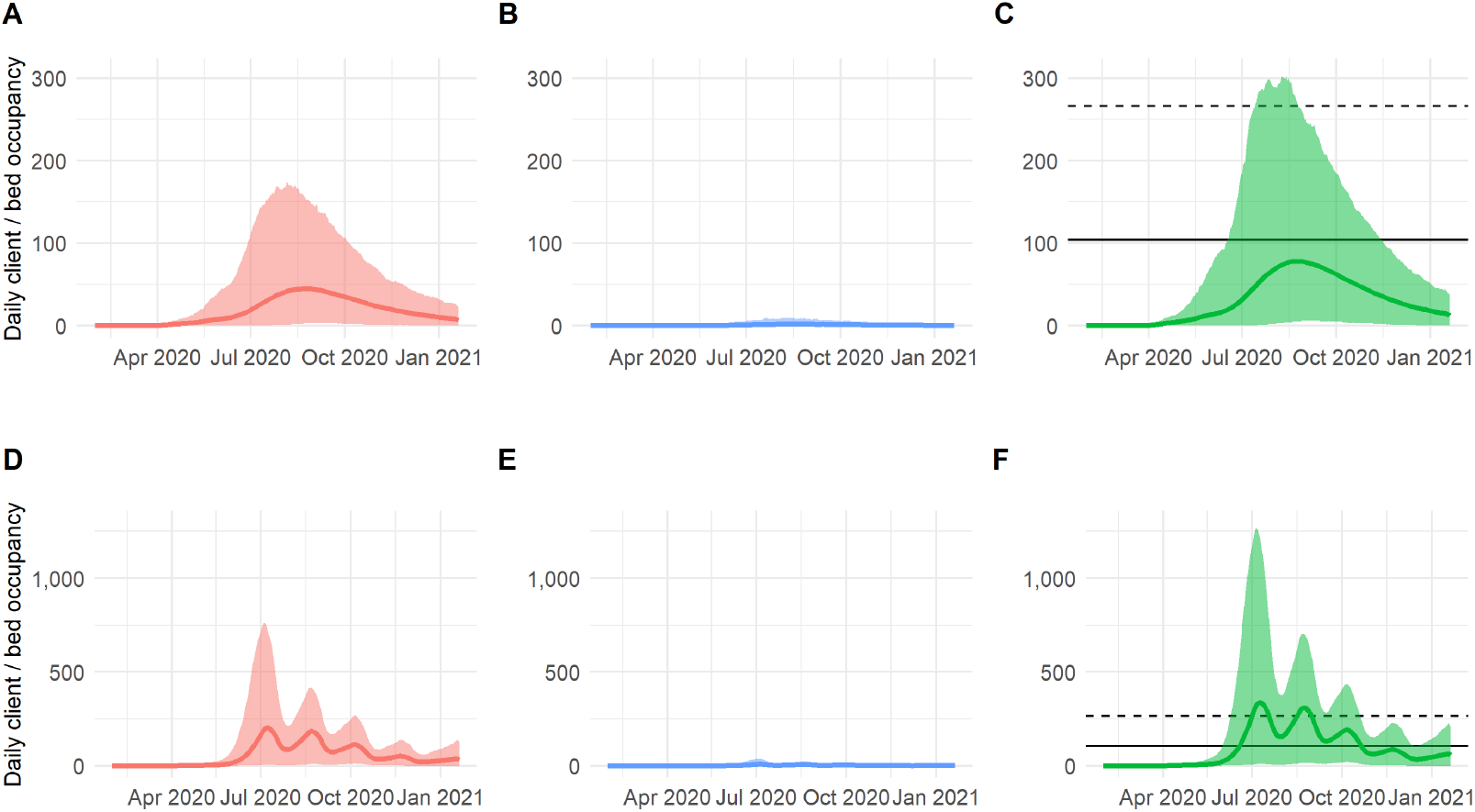
Example post-acute care daily bed occupancy needs and available skilled nursing facility beds in COVID-19 dedicated facilities (solid black line) and beds in COVID-19 dedicated wards within a larger facility (dotted black line) in the State of Utah for patients discharged from the ICU to each of three care types under two non-pharmaceutical interventions A) Home health care based on current Utah, B) Hospice care based on current Utah, and C) Skilled Nursing Facilities based on current Utah, D) Home health care based on pulsed social distancing, E) Hospice care based on pulsed social distancing, and F) Skilled Nursing Facilities based on pulsed social distancing.

**Fig 6.**
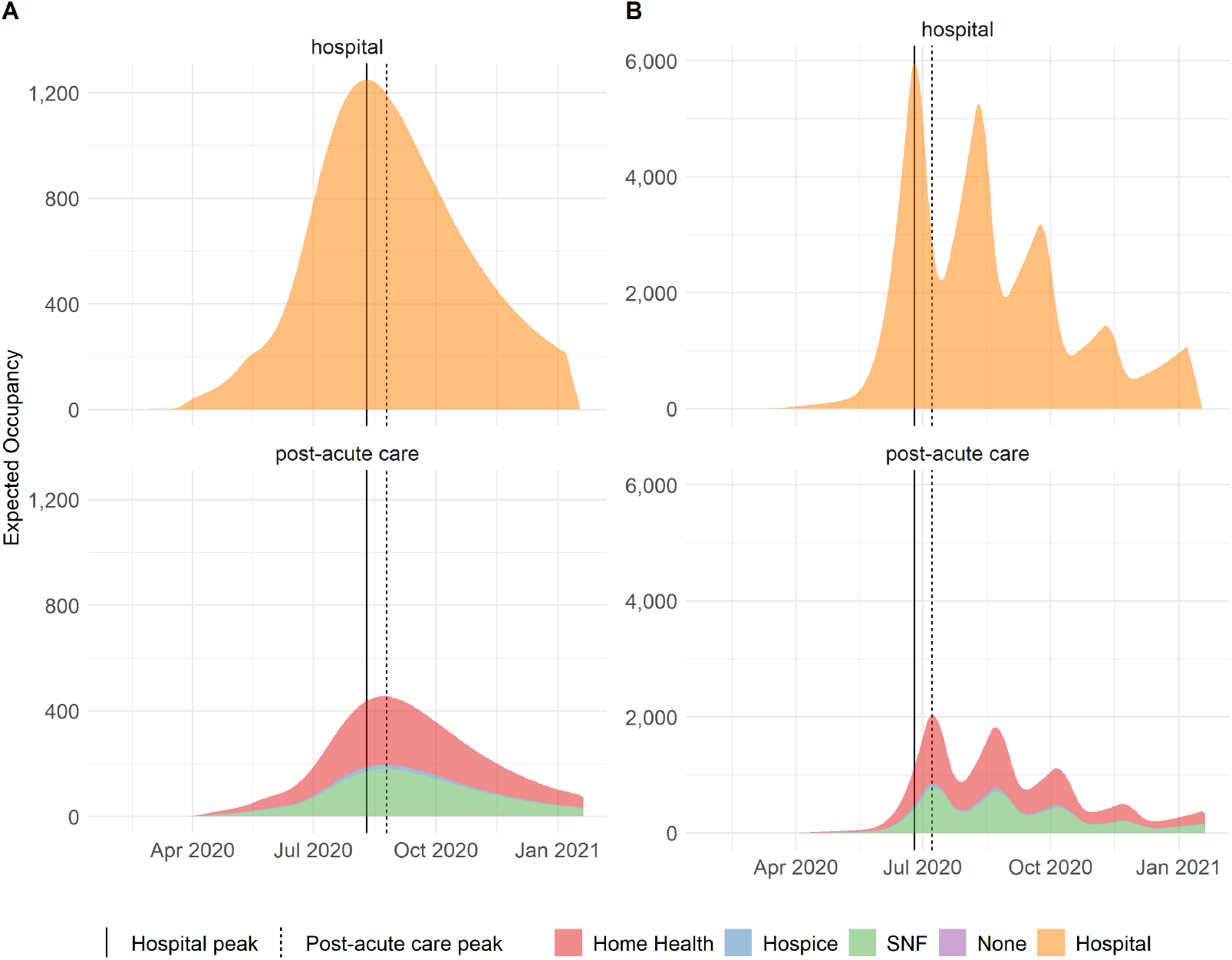
Daily hospital and post-acute care occupancy for (A) current Utah and (B) pulsed social distancing.

Likewise, while the overall shape of the post-acute care occupancy curves are similar for patients discharged from the hospital and from the ICU, we see that a much greater fraction of patients discharged from the ICU requires skilled nursing care (Figure 5) compared to a nearly equal proportion of patients discharged from the hospital requiring home health care and skilled nursing care (Figure 4).

Like most epidemiological models that show acute care hospitalizations and ICU admissions changing under various non-pharmaceutical interventions, we show how our post-acute care model can take in discharge estimates from different outbreak curves, leading to different post-acute care needs at different times (Figures 4 and 5).

## Discussion

We present the COVID-19 post-acute care model as an open-source modeling framework that aims to extend the standard output of most epidemiological models (i.e., hospitalization and ICU discharge) to fill a major gap in modeling the COVID-19 outbreak. The post-acute care model aims to guide public health policymaking in addressing the “aftershocks” of discharged patients requiring further care. Aside from filling a major gap in the literature [10], the flexibility of our model to accept a time series of discharges generated from any epidemiological model to forecast the post-acute care needs of discharged hospital and ICU patients will enable this model to be adapted for a multitude of different outbreak scenarios across multiple spatial and temporal scales.

Although the COVID-19 pandemic is well underway in most locations, with the long duration of stay in hospital and ICU settings, many locations are just now starting to experience the first surge of patients into post-acute care services. Thus, this model represents an important tool in guiding public health decision-making towards how patients will receive prolonged care post-discharge. Arranging bed availability may be especially important as the infection status of these patients may be uncertain [18–20]. Likewise, discharging potentially COVID-19 positive patients to skilled nursing facilities poses a potential problem as evidenced by the significant outbreaks in long term care facilities [11]. Consequently this model can help guide the creation and/or expansion of COVID-specific post-acute care services.

Further, while little information is available on the types of services that patients with COVID-19 need following discharge from acute care, data on post-discharge probability is also still preliminary. Here, we develop the model and implement probabilities based on preliminary data from Utah, however we also highlight the way in which our prior distributions can be updated as new data become available or replaced entirely with data from other locations.

While this model projects the bed needs for different post-acute care services, it is important to consider the organization necessary to accommodate the influx of recovering COVID-19 patients and protect non-COVID-19 patients. Ideally, specialized buildings or units would be repurposed for COVID-19 patients to recover in. However, many facilities currently lack the design and/or staffing resources necessary to isolate recovering or quarantined COVID-19 patients [21]. It is critical for public health officials and post-acute care leaders to work together to determine which facilities have the beds, staff, and resources necessary to support the influx of patients and create protocols for accepting and placing recovering COVID-19 patients while protecting vulnerable patients.

This model can also be used and adapted to explore potential alternative strategies for addressing the surge in acute care facilities during future pandemic waves. As suggested by Grabowski et al. [10], post-acute care services can act as a “pressure release valve” for acute care hospitals, taking in patients who are partially recovered to release beds for more critically ill patients. However, without proper planning, post-acute care facilities can serve as the opposite, acting as a “bottleneck.” If patients who are ready to be discharged but require additional care have nowhere to go as a result of post-acute care services being saturated, this can negatively impact hospital capacity. While exploring this relationship is outside the scope of this paper, this modeling framework can be easily adapted to explore these and simliar questions.

Additionally, as treatments for COVID-19 are developed, one unanticipated side effect could be that while treatments successfully prevent death in a large number of patients, they could result in more patients, patients who would have died without treatment, surviving and requiring long-term care. At the current time, any estimates of how treatments may impact acute or post-acute care needs would be entirely speculative, however, as more becomes known about potential successful treatments, the prior estimates for proportion of patients being discharged to the different care services can easily be updated to adapt to this type of new information.

Our modeling approach has several limitations. Our initial assumptions about the fraction of patients requiring each post-acute care service are very uncertain, as they are based on aggregated historical University of Utah Health discharges prior to the appearance of COVID-19. However, we account for this by giving relatively low weight to our priors versus observed COVID-19 discharges. Another limitation is that we do not account for uncertainty about the length-of-stay estimates for post-acute care services, although the model could be modified to incorporate this in future work. The largest limitation is that the model does not directly account for characteristics of the patient population, meaning the posterior allocation probabilities may not generalize well to regions or health systems with dissimilar populations. An extension of this work could condition the probability of requiring a post-acute care service on patient characteristics such as age or comorbidities. Although we highlight several limitations, our approach is well-suited to answer policy questions and provides a unique modeling output that can help guide the post-acute care surge. As most epidemiological models focused on acute care services, we concentrate on post-discharge needs, however, as is the case with standard epidemiological models, it is critical for policy makers to consider the range of possible trajectories and the sensitivity of results to different assumptions. Likewise, since we do not model the community outbreak leading to hospitalizations, it is critical that the inputs to our post-acute care model be carefully considered for their strengths and weaknesses, as any limitations of the input model will flow through our model framework as well. Despite these limitations, this model has already been used to help improve post-acute care services in Utah and we believe its usefulness in pandemic preparedness extends far beyond the state.

## Data Availability

Data are available in our Github repository "mattrmaloney/covid-post-acute-care"

https://github.com/mattrmaloney/covid-post-acute-care

## Author Contributions

Conception and design of study: MM, RM, RC, PW, DB Acquisition of data: RC, PW, MGH, HRM, LTK Development and/or verification of analytic methods: MM, RM, RC, DB, LTK Analysis and/or interpretation of data: MM, RM, RC, DB, MGH, MHS, LTK Drafting the manuscript: MM, RM, LTK Revising the manuscript: MM, RM, RC, PW, DB, HRM, MGH, MHS, LTK Approval of the final manuscript: MM, RM, RC, PW, DB, HRM, MGH, MHS, LTK

## References

[1] Zhu N, Zhang D, Wang W, Li X, Yang B, Song J, et al. A novel coronavirus from patients with pneumonia in china, 2019. New England Journal of Medicine 2020.

[2] Dong E, Du H, Gardner L. An interactive web-based dashboard to track covid-19 in real time. The Lancet Infectious Diseases 2020.

[3] Huang C, Wang Y, Li X, Ren L, Zhao J, Hu Y, et al. Clinical features of patients infected with 2019 novel coronavirus in wuhan, china. The Lancet 2020;395:497–506.

[4] Garg S. Hospitalization rates and characteristics of patients hospitalized with laboratory-confirmed coronavirus disease 2019—covid-net, 14 states, march 1–30, 2020. MMWR Morbidity and Mortality Weekly Report 2020;69.

[5] Moghadas SM, Shoukat A, Fitzpatrick MC, Wells CR, Sah P, Pandey A, et al. Projecting hospital utilization during the covid-19 outbreaks in the united states. Proceedings of the National Academy of Sciences 2020;117:9122–6.

[6] Ferguson N, Laydon D, Nedjati Gilani G, Imai N, Ainslie K, Baguelin M, et al. Report 9: Impact of non-pharmaceutical interventions (npis) to reduce covid19 mortality and healthcare demand 2020.

[7] Branas CC, Rundle A, Pei S, Yang W, Carr BG, Sims S, et al. Flattening the curve before it flattens us: Hospital critical care capacity limits and mortality from novel coronavirus (sars-cov2) cases in us counties. medRxiv 2020.

[8] Joseph C. Lemaitre JK Kyra H. Grantz. A scenario modeling pipeline for COVID-19 emergency planning. medRxiv 2020.

[9] Murray C, others. Forecasting the impact of the first wave of the covid-19 pandemic on hospital demand and deaths for the usa and european economic area countries 2020.

[10] Grabowski DC, Maddox KEJ. Postacute care preparedness for covid-19: Thinking ahead. JAMA 2020;323:2007–8.

[11] Comas-Herrera A, Zalakain J, Litwin C, Hsu AT, Lane N, Fernandez J-L. Mortality associated with covid-19 outbreaks in care homes: Early international evidence. International Long Term Care Policy Network 2020.

[12] Commission MPA. Report to congress: Medicare payment policy—skilled nursing facility services. News release 2019.

[13] Iacobucci G. Covid-19: Care home deaths in england and wales double in four weeks 2020.

[14] Condon B, Peltz J, Mustian J. AP count: Over 4,500 virus patients sent to ny nursing homes 2020.

[15] Maloney MR, Checketts R, Keegan LT. mattrmaloney/covid-post-acute-care: First release of COVID post acute care project. Zenodo; 2020. https://doi.org/10.5281/zenodo.3891761.

[16] Zhang Y, Keegan LT, Yuqing Q, Samore MH. The real time effective reproductive number for covid-19 in the united states. medRxiv 2020.

[17] Jarvis CI, Van Zandvoort K, Gimma A, Prem K, Klepac P, Rubin GJ, et al. Quantifying the impact of physical distance measures on the transmission of covid-19 in the uk. BMC Medicine 2020;18:1–10.

[18] Zhang J-f, Yan K, Ye H-h, Lin J, Zheng J-j, Cai T. SARS-cov-2 turned positive in a discharged patient with covid-19 arouses concern regarding the present standard for discharge. International Journal of Infectious Diseases 2020.

[19] Yuan J, Kou S, Liang Y, Zeng J, Pan Y, Liu L. PCR assays turned positive in 25 discharged covid-19 patients. Clinical Infectious Diseases 2020.

[20] Xing Y, Mo P, Xiao Y, Zhao O, Zhang Y, Wang F. Post-discharge surveillance and positive virus detection in two medical staff recovered from coronavirus disease 2019 (covid-19), china, january to february 2020. Eurosurveillance 2020;25:2000191.

[21] Tumlinson A, Altman W, Glaudemans J, Gleckman H, Grabowski DC. Post-acute care preparedness in a covid-19 world. Journal of the American Geriatrics Society 2020.

